# Impact of 10-valent pneumococcal conjugate vaccine on the epidemiology of otitis media with otorrhea among Bangladeshi children

**DOI:** 10.1101/2024.09.21.24314121

**Authors:** Hakka Naziat, Md Hafizur Rahman, Rajib C Das, Maksuda Islam, Shampa Saha, Md. Manzoor Hussain, Asif Sattar, Mohammad J Uddin, Md. Jahangir Alam, Cynthia G. Whitney, Senjuti Saha, Yogesh Hooda, Samir K Saha

## Abstract

In 2015, Bangladesh introduced the ten-valent pneumococcal conjugate vaccine (PCV10). This study evaluates the impact of PCV10 on pneumococcal otitis media (OM), a prevalent middle ear infection amongst children in low-and-middle income countries, including Bangladesh. We analyzed ear swabs from OM cases with otorrhea collected at the largest pediatric hospital in Dhaka, Bangladesh from April 2014 to March 2019. The study involved identifying pathogens and conducting pneumococcal serotyping using the Quellung reaction.

Four years post PCV10 introduction, the rate of detection of pneumococcus among otorrhea cases decreased from 18.4% (164/892) in 2014-15 to 15.6% (581/3735) in 2018-19, reflecting a 15.4% reduction. Notably, vaccine serotypes (VTs) demonstrated a significant decline throughout the post-PCV years, with a 44.1% reduction compared to the pre-PCV period. Specific VTs like 14, 6B and 19F exhibited significant reductions of 60.2%, 57.5% and 42.9% respectively. Conversely, non-vaccine serotypes (NVTs) showed non-significant increase of 8.6%; specifically serotype 35B showed a 70.1% increase. Administration of two or three doses of PCV10 provided 84.4% protection against OM cases caused by serotypes covered by PCV10.

This is the first report from South Asia assessing PCV’s impact on pneumococcal OM. It demonstrates a 15.4% reduction in pneumococcal isolation over four years and over 80% efficacy of PCV10 in preventing vaccine type pneumococcal OM cases. While some vaccine serotypes decreased significantly, the impact of PCV10 on overall pneumococcal OM cases was dampened by increasing isolation of non-vaccine serotypes.

## INTRODUCTION

*Streptococcus pneumoniae*, or pneumococcus, is a significant bacterial pathogen responsible for a range of pediatric infections worldwide, including meningitis, sepsis, pneumonia and otorrhea [1,2]. Given the considerable global burden of pneumococcus-associated morbidity and mortality, the World Health Organization has recommended the inclusion of pneumococcal conjugate vaccine (PCV) in the routine immunization programs of all countries and assessment of its impact on pneumonia, other invasive diseases, and otitis media (OM) [3,4]. In March 2015, Bangladesh introduced the 10-valent pneumococcal conjugate vaccine (PCV10), targeting 10 serotypes: 1, 4, 5, 6B, 7F, 9V, 14, 18C, 19F, and 23F [5]. In addition, PCV10 was also anticipated to provide cross-protection against 6A due to immunologic similarities with vaccine serotype 6B [6–8]. Bangladesh has successfully achieved high coverage (99.2% coverage for ≥ 2 doses) of PCV10 across the country [9], and recent studies have reported a decline in severe radiologically confirmed pneumonia amongst children under the age of five years at two sites in rural Bangladesh – Mirzapur [5] and Sylhet [10]. Nevertheless, the broader impact of PCV10 on other pneumococcal diseases remain to be assessed.

Otitis media is among the most common bacterial infections diagnosed in children globally, with peak incidences typically within the first two years of life [11,12]. OM can range from asymptomatic cases, which usually resolve or are treated without complications, to more severe forms such as recurrent, nonresponsive, dry perforation, spontaneously draining, and chronic suppurative OM [13]. In Bangladesh, pneumococcus is a principal cause of OM, accounting for 18% of all OM cases [14]. PCV10 and PCV10 + 6A serotypes accounted for 8% and 9% of all OM cases and 46% and 49% of pneumococcus-positive cases, respectively, and these serotypes exhibited a higher likelihood to be non-susceptible to at least one antibiotic (erythromycin and/or trimethoprim-sulfamethoxazole) than nonvaccine serotypes (91% vs. 77%). A recent Cochrane review including 15 publications from 11 trials reported that administration of the licensed seven-valent pneumococcal conjugate vaccine (PCV7) and ten-valent PCV (PCV10) during early infancy substantially reduced the relative risk of pneumococcal OM [15].

To assess the impact of PCV10 on pneumococcal OM, our research group conducted a study to identify and characterize bacterial pathogens of OM in the ear, nose, and throat (ENT) outpatient department of Bangladesh Shishu Hospital and Institute from 2014 to 2019, spanning one year prior to PCV10 introduction and four years following the PCV10 introduction. The study aimed to determine a) the impact of PCV10 on percentage of OM cases that were pneumococcus-positive, b) changes in serotype distribution amongst pneumococcal OM cases, c) variations in the age of infection and duration of ear discharge, and d) the antimicrobial susceptibility of pneumococcus isolated from OM cases.

## METHODS

The methodology employed in this study adhered strictly to those outlined in our previously published baseline study [14], with no modifications. This consistency ensured the comparability of results across both study periods. Details are provided below.

### Study site and population

The study was conducted at Bangladesh Shishu Hospital and Institute (BSHI, formerly known as Dhaka Shishu Hospital or DSH), the largest pediatric hospital in Bangladesh, which serves children from 0 to 18 years. This hospital has a dedicated out-patient department (OPD) for the management of all cases seeking care for ear, nose, and throat (ENT) conditions.

### Case identification and enrollment criteria

Children presenting with symptoms suggestive of otitis media (OM) were identified by a study physician based on World Health Organization-recommended clinical criteria and otoscopic examinations [16,17]. The inclusion criteria for OM cases were: (1) acute onset of one or more sign/symptoms of infection such as fever, earache, ear tugging or irritability, and (2) middle ear inflammation with/without middle ear effusion or spontaneous discharge (otorrhea). From enrolled children with/without otorrhea (not caused by otitis externa), data on sex, age, duration of discharge, and antibiotic use within the previous seven days were recorded. Informed consent was obtained from caregivers for children with otorrhea to collect specimens.

### Specimen collection

Specimens were collected from children presenting with otorrhea by trained research assistants. The external auditory canal was swabbed after the removal of any debris, using a flocked swab, and placed into a tube containing STGG medium (2% skim milk, 3% tryptone, 10% glycerol and 0.5% glucose). Each specimen was cultured within three hours in the laboratory and examined for the presence of bacterial pathogens.

### Laboratory methods

All specimens were cultured on blood agar with and without gentamicin (in 5% CO_2_ environment), chocolate agar with bacitracin and MacConkey agar [17]. Growth on these selective media were examined for the presence of *Haemophilus influenzae*, *S. pneumoniae*, and other bacterial pathogens, such as *Staphylococcus aureus*, *Moraxella catarrhalis*, *Pseudomonas spp*.

Pneumococcal isolates were serotyped using the Quellung reaction [18]. Antibiotic susceptibility testing was conducted using the disc diffusion method adhering to the latest Clinical and Laboratory Standards Institute (CLSI) guidelines [19]. Antibiotics tested included penicillin (1 μg oxacillin disc), trimethoprim-sulfamethoxazole (1/19, 25 μg disc), chloramphenicol (30 μg disc), erythromycin (15 μg disc) and ciprofloxacin (5 μg disc). Non-susceptible pneumococcal isolates were further tested using the E-test to determine minimum inhibitory concentrations.

### Indirect cohort analysis

Vaccine effectiveness was assessed by comparing the vaccination status of children with vaccine-type OM (VT-OM) and non-vaccine-type OM (NVT-OM). This analysis assumes that vaccination does not influence the risk for non-vaccine-type disease among vaccinated individuals. A recent study has shown that results derived from the indirect cohort method were comparable to that of a standard case-control study [20,5], which is complex and difficult to implement. VT-OM was defined as OM cases in which PCV10 serotypes or 6A were identified, all other serotypes were considered NVT. All pneumococcal OM cases from April 2015 through March 2019 were used for this analysis. Based on the immunization status verified by the vaccination card or the caregiver’s verbal confirmation (in cases where the card was not available or lost), children were classified into two groups: vaccinated and unvaccinated. The unvaccinated cohort included verbally confirmed non-vaccinated children and 2 - <24-month-old children who were not age-eligible for PCV at the time of the initial pneumococcal vaccination program (21st March 2015). As Bangladesh’s EPI did not have catch-up campaigns and only children aged 6 weeks were eligible for vaccine introduction, this strategy led to an increase of vaccine eligible children gradually with time. To identify vaccine eligible children, we back calculated the birthdate of children and considered those born on 7th February 2015 or later as eligible to receive the 1st dose of pneumococcal vaccine at 6 weeks of age. Children who sought medical care two weeks or more after receiving an initial dose were considered vaccinated. Any child who came to the hospital within 14 days of receiving the 1st dose of PCV10 vaccine was considered non-vaccinated. Children who received two or three doses of PCV10 were considered individually for calculating vaccine impact, using children with no doses were used the comparison group. We calculated the odds ratio of receipt of PCV10 dose among children who had vaccine-type (PCV10) OM versus non-vaccine-type OM and used logistic regression to estimate vaccine effectiveness, calculated as: Vaccine effectiveness = (1-odds ratio for PCV10 vaccination) x 100%.

### Statistical data analysis

Data analyses were conducted using STATA 14.0 (Stata Corp LP, College Station, TX). Epidemiological data were analyzed to elucidate the serotype distribution amongst pneumococcal OM cases, duration of ear discharge, and antibiotic susceptibility profiles of *S. pneumoniae* isolates. Common contaminants (e.g., *Bacillus spp.*, *Staphylococcus spp.* etc.) were excluded.

### Ethical clearance

This surveillance protocol was approved by the Institutional Review Board of Bangladesh Institute of Child Health at BSHI. Specimens were collected as part of routine patient care.

## RESULTS

From April 2014 to March 2019, a total of 5,040 OM cases were identified, with 4,279 (93.8%) classified as otorrhea cases (Figure 1, Table 1). Ear swabs were successfully collected from 4,626 (97.8%) of these cases. A pathogen could be identified in 2,181 of the 4,626 samples (47.1%; Supplemental Table 1). *H. influenzae* was the most frequently detected pathogen (782/2,181; 35.8%), followed by *S. pneumoniae* (581/2,181; 26.6%), *S. aureus* (514/2,181; 23.6%) and *Pseudomonas spp.* (365/2,181; 16.7%). The trends in isolation of all detected causes of OM cases are shown in Figure 2. In 554 out of the 2,181 culture-positive cases (25.4%), multiple organisms were detected.

**Figure 1:**
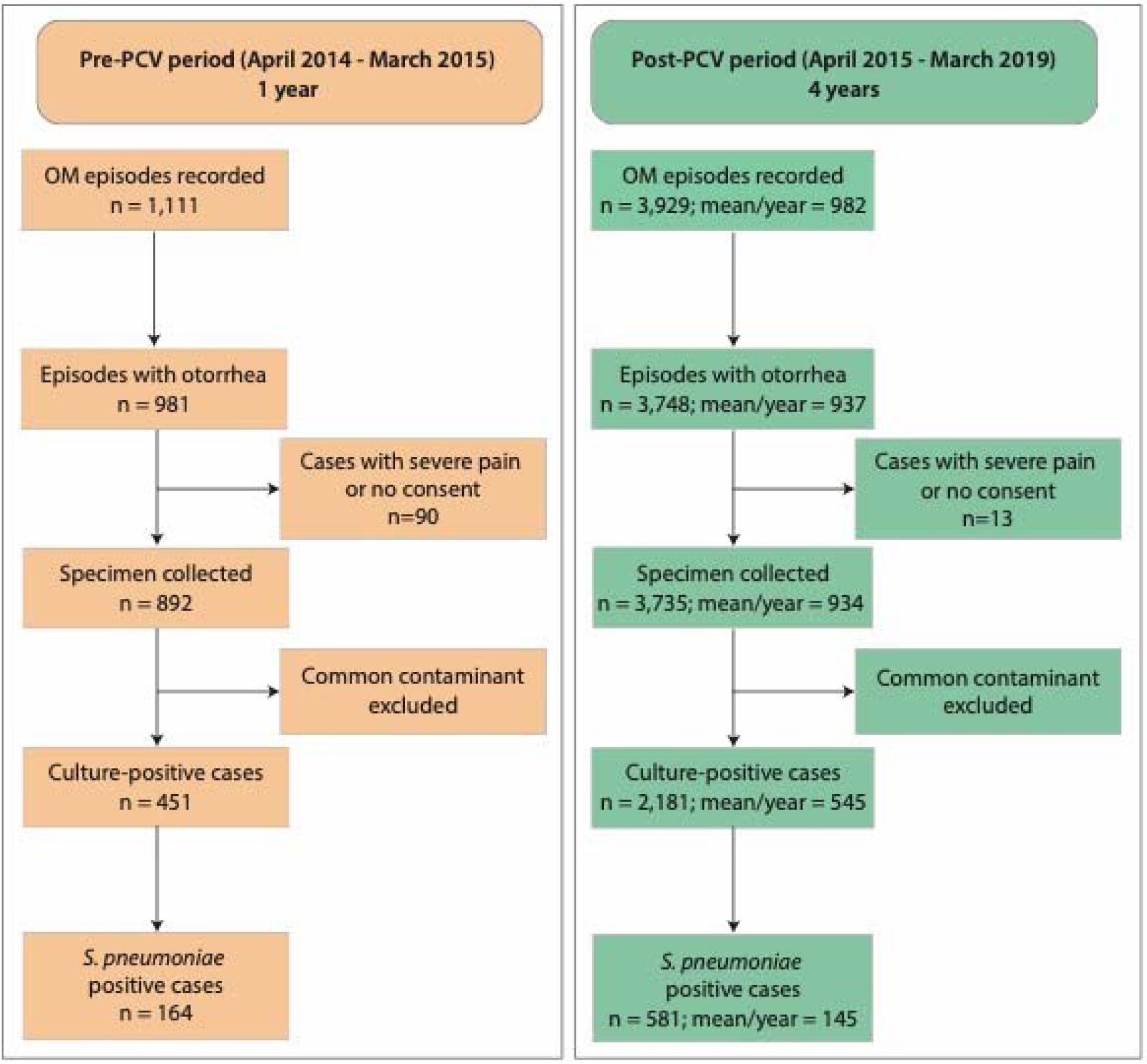
Study design highlighting the number of yearly *S. pneumoniae* isolates collected during the baseline (Pre-PCV, April 2014-March 2015) and present study period (Post-PCV, April 2015-March 2019).

**Table 1:**
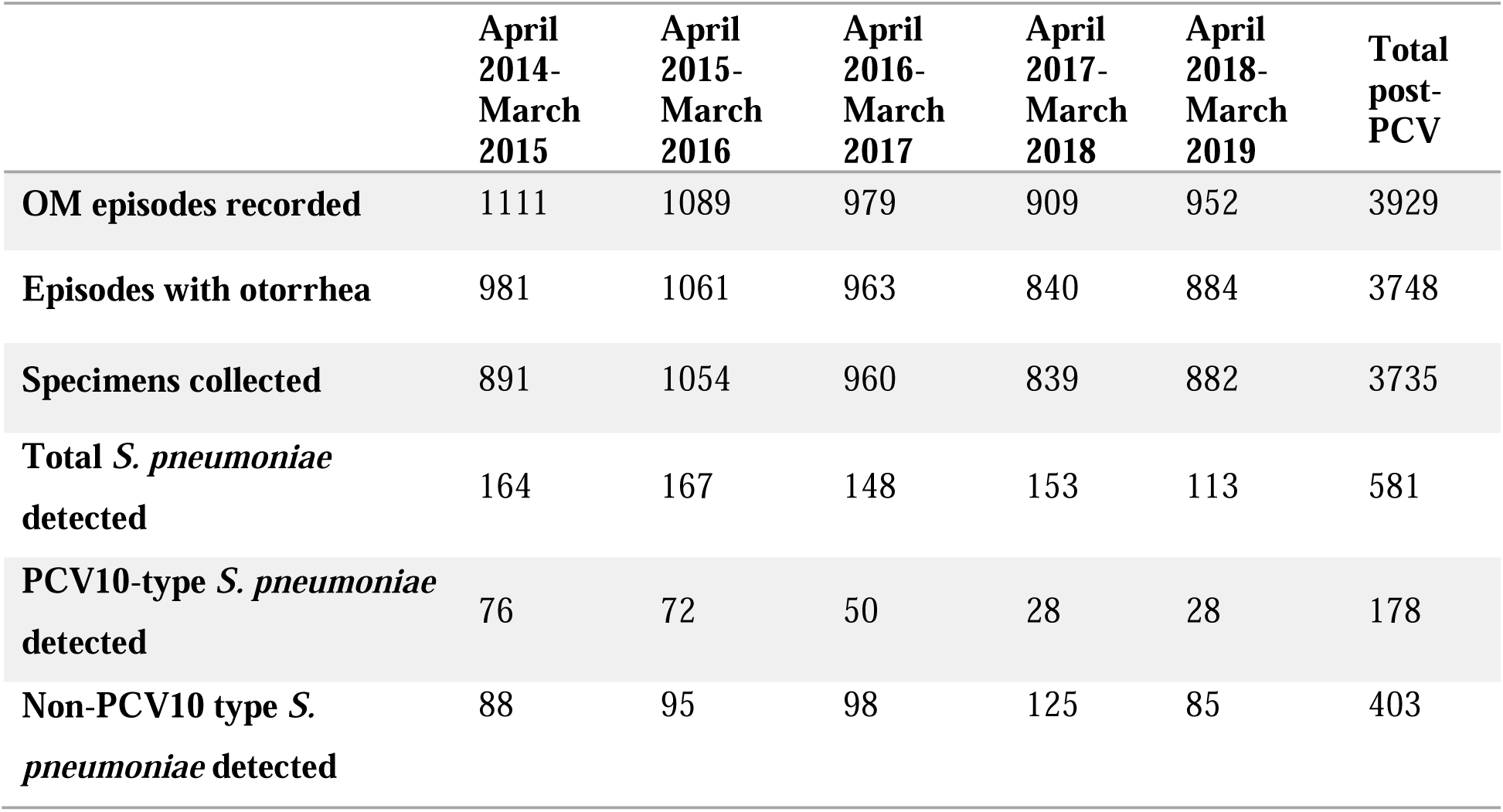
Total, PCV10-type and non-PCV10-type *S. pneumoniae* obtained from OM cases in this study.

### Trends in number and serotype distribution of pneumococcal positive otorrhea cases

From April 2014 to March 2015, 164 pneumococcal OM cases were identified. This number reduced to 113 by April 2018 to March 2019. Over the four years following the introduction of PCV10, the pneumococcal positivity among otorrhea cases decreased from 18.4% (164/892) to 15.6% (581/3,735) [15.4% reduction (95% CI: −1.2-28.9%); p=0.04] (Figure 3). The detection rate for *Haemophilus influenzae, Staphylococcus aureus and Pseudomonas spp* was not affected (Figure 2).

**Figure 2:**
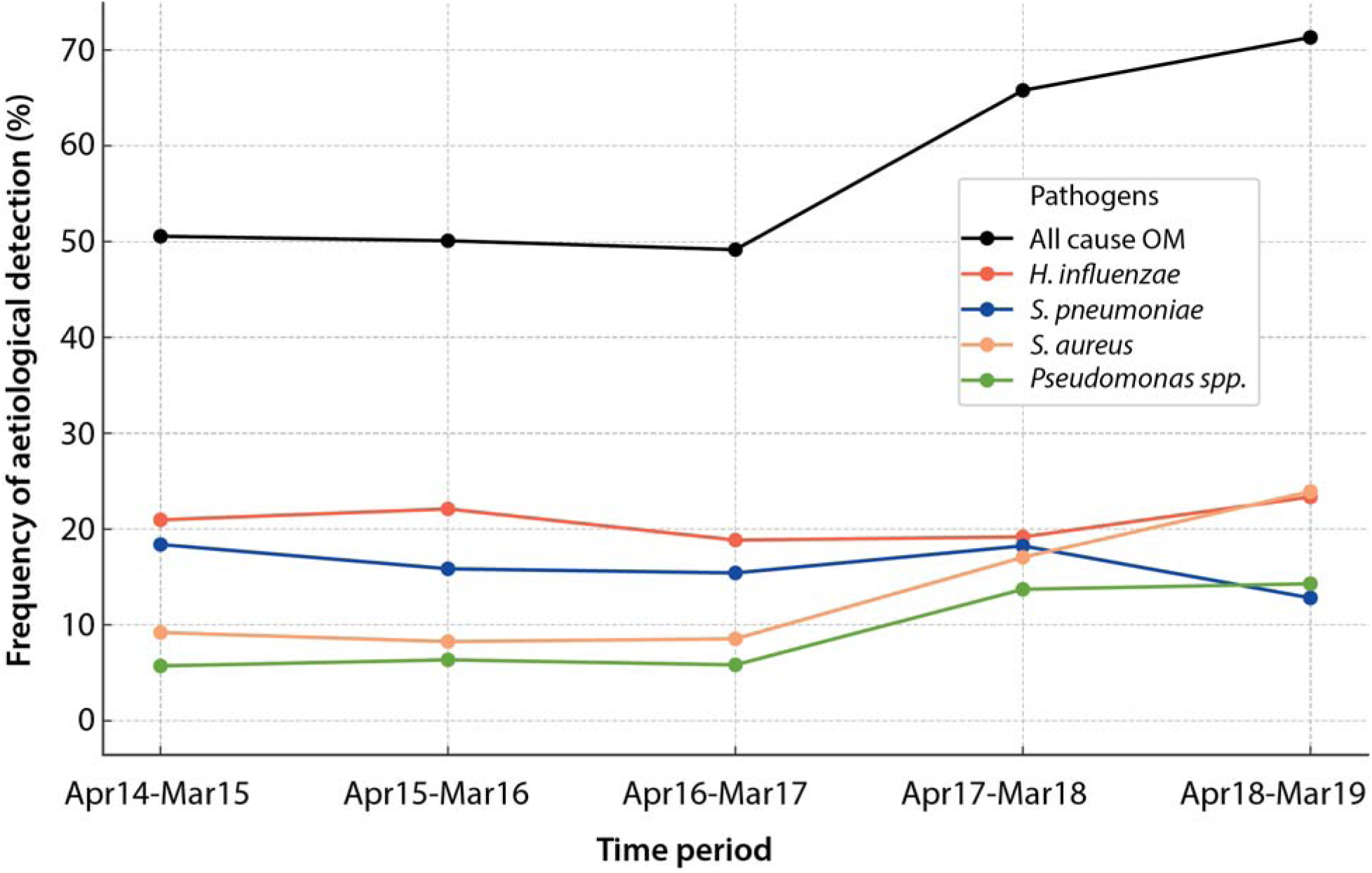
Trends in isolation of all cause OM, and OM caused by *H. influenzae*, *S. pneumoniae*, *S. aureus*, *Pseudomonas spp*.

Vaccine serotypes displayed a declining trend over the post-PCV year, resulting in a 44.1% (95% CI: 25.8 - 57.5%, p<0.001) reduction compared to the pre-PCV period (Figure 3). Notably VTs like 14, 6B and 19F showed significant reductions of 60.2% (95% CI: 10.7% - 81.7%; p=0.009), 57.5% (95% CI: −9.01-82.3%; p=0.026) and 42.9% (95% CI: 2.6-65.5%; p=0.015), respectively (Figure 4A). No protection was seen for Serotype 1 (35.6%, 95% CI: −85.7%-83.7%; p=0.215). While the percentage of OM cases attributable to 6A declined, this change was not statistically significant (10.4% reduction, 95% CI: −270.7% - 71.5%; p=0.405).

Non-PCV10 serotypes showed a slight increase although not significant (8.6% increase, 95% CI: −15.4-28.2%, p=0.225), particularly 35B, which exhibited a 70.1% increase (95% CI: −18.6 % - 96.6%; p=0.04) (Figure 3, Sup Figure 1). Certain serotypes absent in the pre-PCV period were notably present in substantial numbers during the post-PCV period, including serotype 17F (N=13), 12F (N=9), and 18A (N=7) (Sup Figure 1).

**Figure 3:**
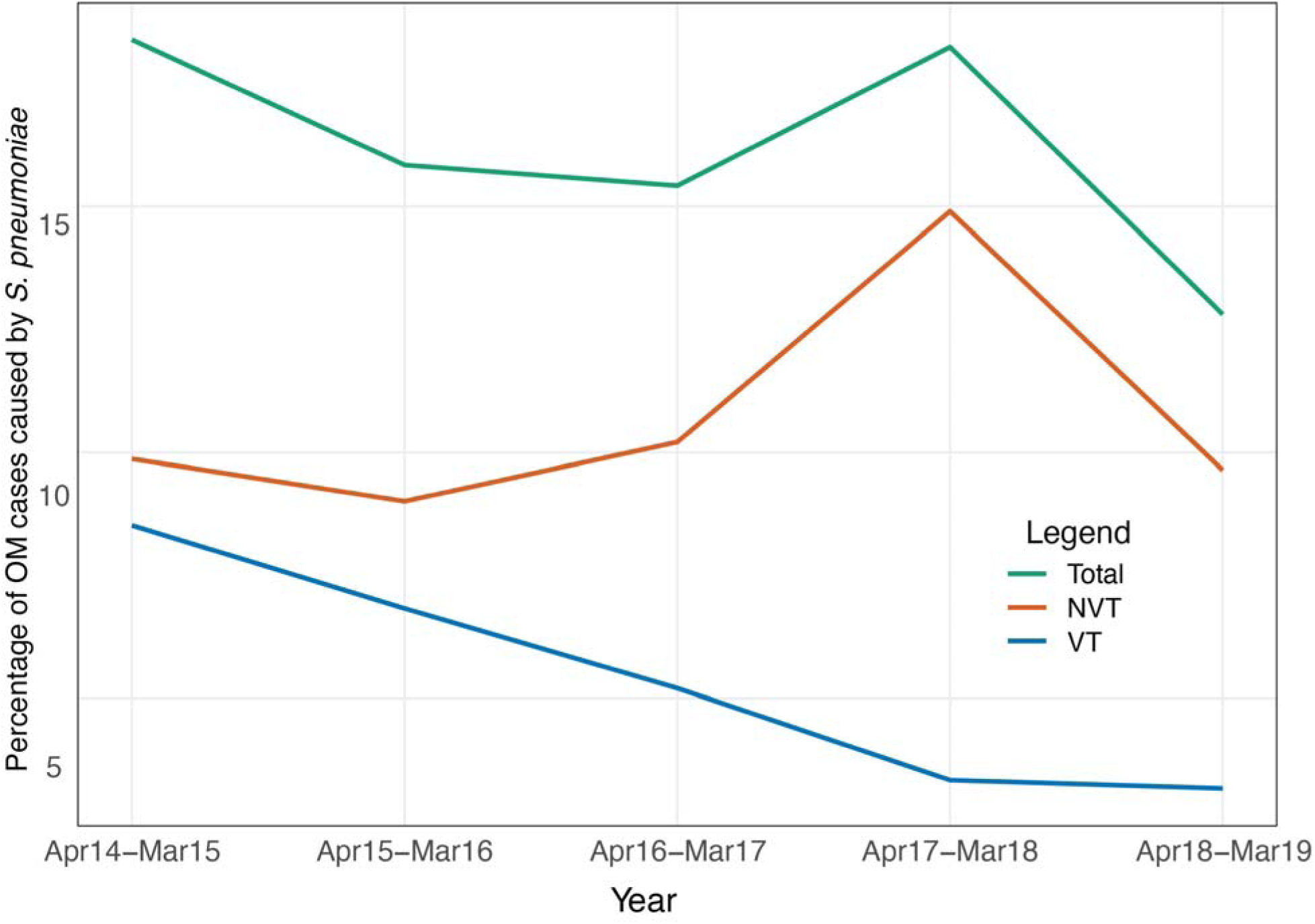
Percent of OM cases attributable to *S. pneumoniae* at Bangladesh Shishu Hospital and Institute in Dhaka, Bangladesh from April 2014 to March 2019. Trends of all *S. pneumoniae* (green), PCV10-type *S. pneumoniae* (VT, blue) and non-PCV10-type *S. pnuemoniae* (NVT, red) isolated from OM cases is shown.

**Figure 4:**
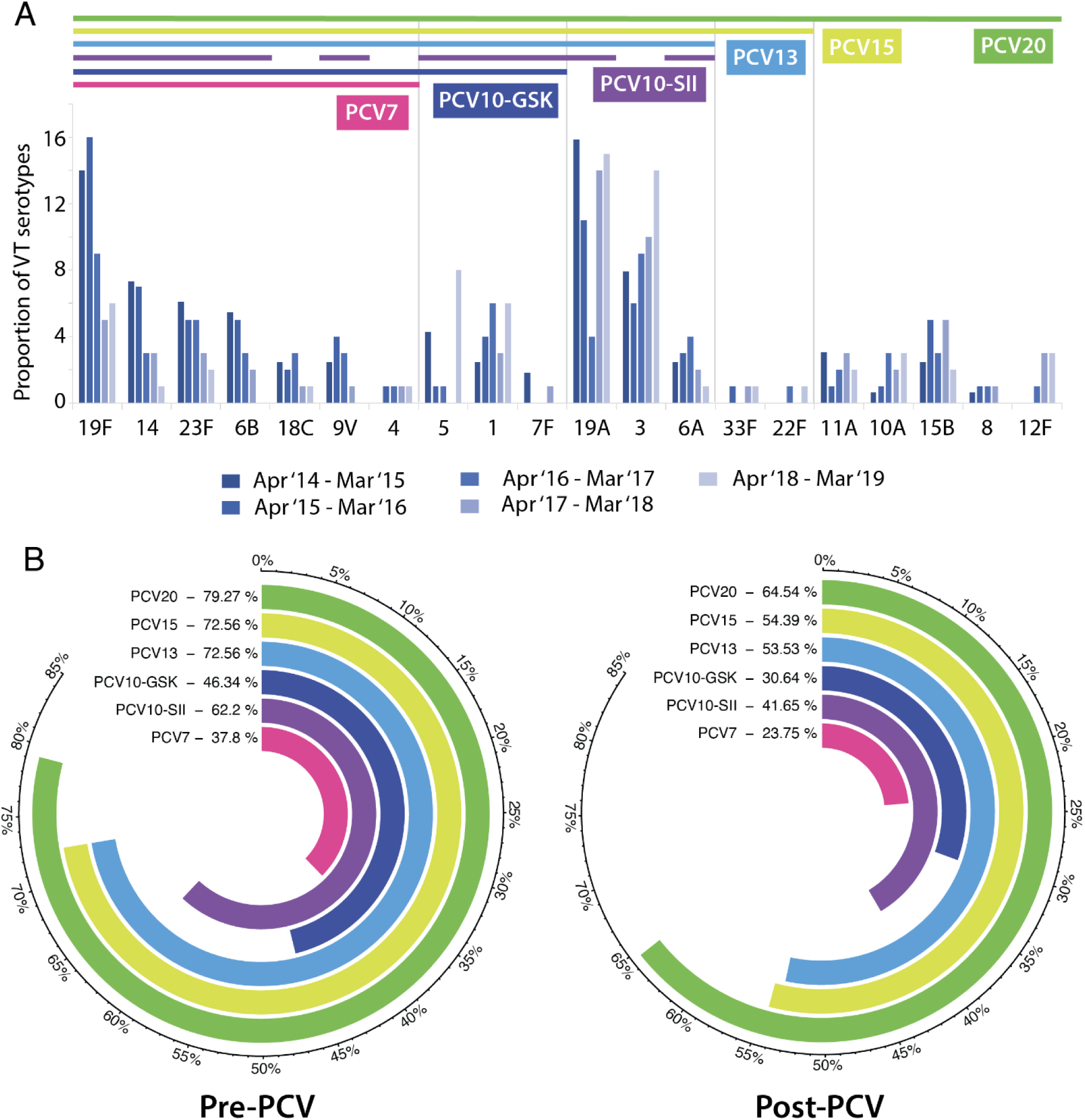
Trends in the proportion of OM cases attributable to vaccine-type *S. pneumoniae* from April 2014 to March 2019. A) The yearly trends of percentage of VT-OM cases caused by different VT pneumococcal serotypes included in PCV7 (pink), PCV10-GSK (dark blue), PCV10-SII (purple), PCV13 (sky blue), PCV15 (yellow) and PCV20 (green) are shown. B) The estimated coverage of different PCV vaccines in the pre-PCV and post-PCV period.

Based on our data, the estimated serotype coverage of different PCVs for OM cases amongst Bangladeshi infants during the pre-PCV period (2014-2015) was 37.8% for PCV7, 46.3% for PCV10_GSK_ 62.2% for PCV10_SII_, 72.6% for PCV13, 72.6 % for PCV15 and 79.3% for PCV20. Four years after the introduction of PCV10 (2018-2019), the estimated serotype coverage changed to 23.8% for PCV7, 30.6% for PCV10_GSK_, 41.7% for PCV10_SII_, 53.5% for PCV13, 54.4% for PCV15 and 64.5% for PCV20 (Figure 4B).

### Age and ear discharge duration

The median age of a child with pneumococcal OM was 10.0 months (95%CI: 9.0-13.0) in the pre-PCV period and 13.0 months (95%CI, 11.0-14.0) in the post-PCV period (Sup Figure 2). The median ear discharge duration was 5 days (95% CI: 5-6) in pre-PCV period and 5 days (95% CI: 4-5) in the post-PCV period (Sup Figure 3). Trend analysis found no statistical difference in the median age or ear discharge duration throughout the study period.

### Trends of antibiotic susceptibility of pneumococcus

A large percentage of pneumococcal isolates were resistant to cotrimoxazole (58%) and erythromycin (72%), while low rates of resistance were observed for penicillin (0.95%), chloramphenicol (5.5%) and ciprofloxacin (6.9%). No statistical difference in trends was observed for the five antibiotics tested over the study period (Figure 4).

**Figure 4:**
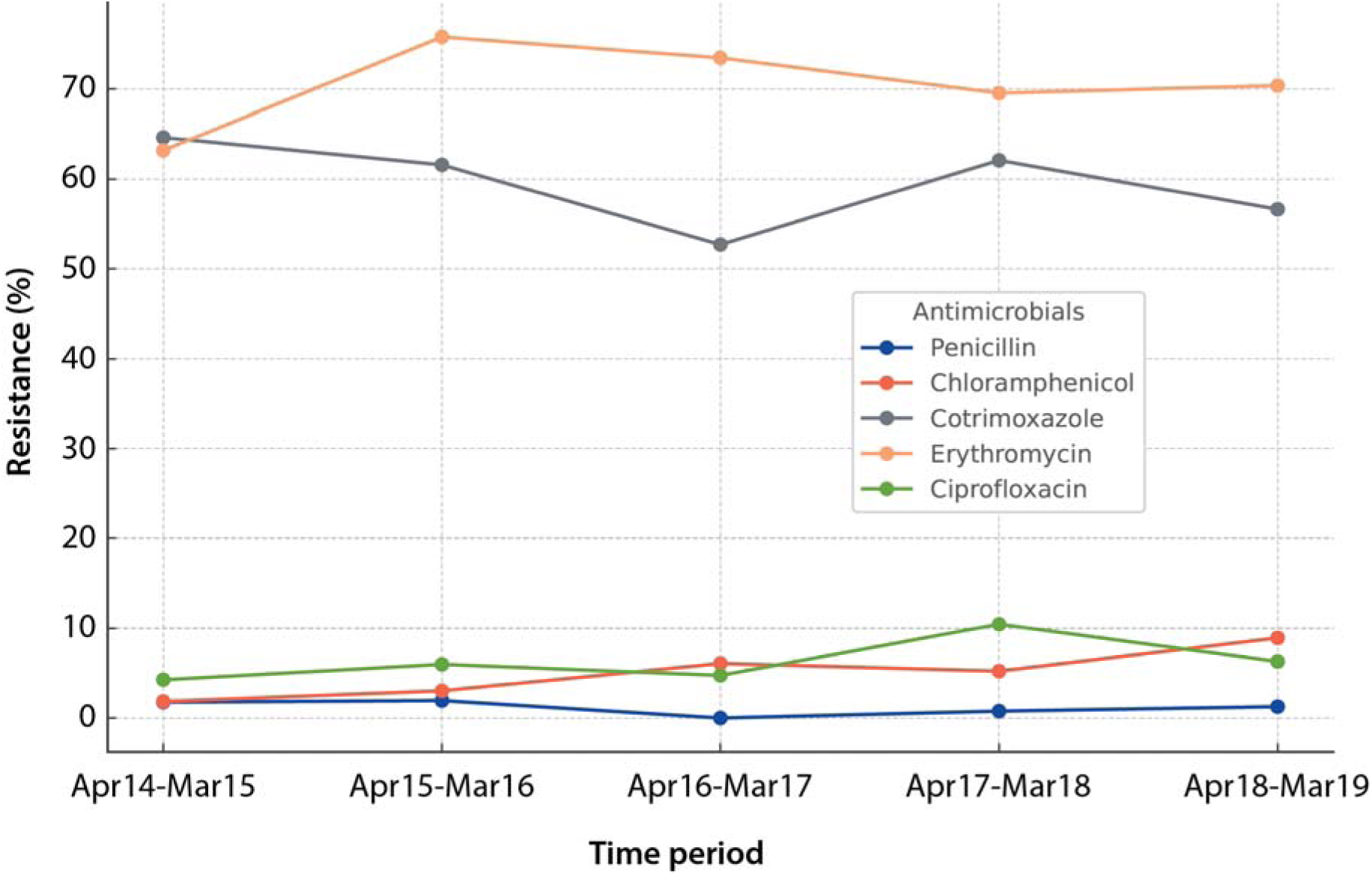
Trends in antimicrobial susceptibility of *S. pneumoniae* isolates against five antibiotics (penicillin, chloramphenicol, cotrimoxazole, erythromycin and ciprofloxacin the during the study period. The proportion of isolates that were resistant to each drug is shown.

### PCV10 effectiveness calculation

Between March 21, 2015, and March 31, 2019, 581 cases of pneumococcal OM were identified, with 323 children being eligible for PCV10 vaccination based on their age. Among these, 178 (30.6%) were of vaccine-type (VT) and 403 (69.4%) were non-vaccine-type (NVT). Of the vaccinated children (n=307), 55 had VT OM, and 252 had NVT OM. Among the unvaccinated (n=274), 123 had VT OM, and 151 had NVT OM. The median age of vaccinated children and unvaccinated children were 7.0 and 33.5 months, respectively. The effectiveness of the 1^st^ PCV10 dose was 83.5% (95% CI: 29.4% - 96.2%; p-value =0.011), 2^nd^ PCV10 dose was 82.1% (95% CI: 42.2% - 94.5%; p-value =0.003) and 3^rd^ PCV10 dose was 84.3% (95% CI: 57.1% - 94.5%; p-value<0.001) (Table 2). Two or three PCV10 doses provided 84.4% (95% CI: 57.1% – 94.3%; p-value <0.001) protection against VT OM (Table 2).

**Table 2:**
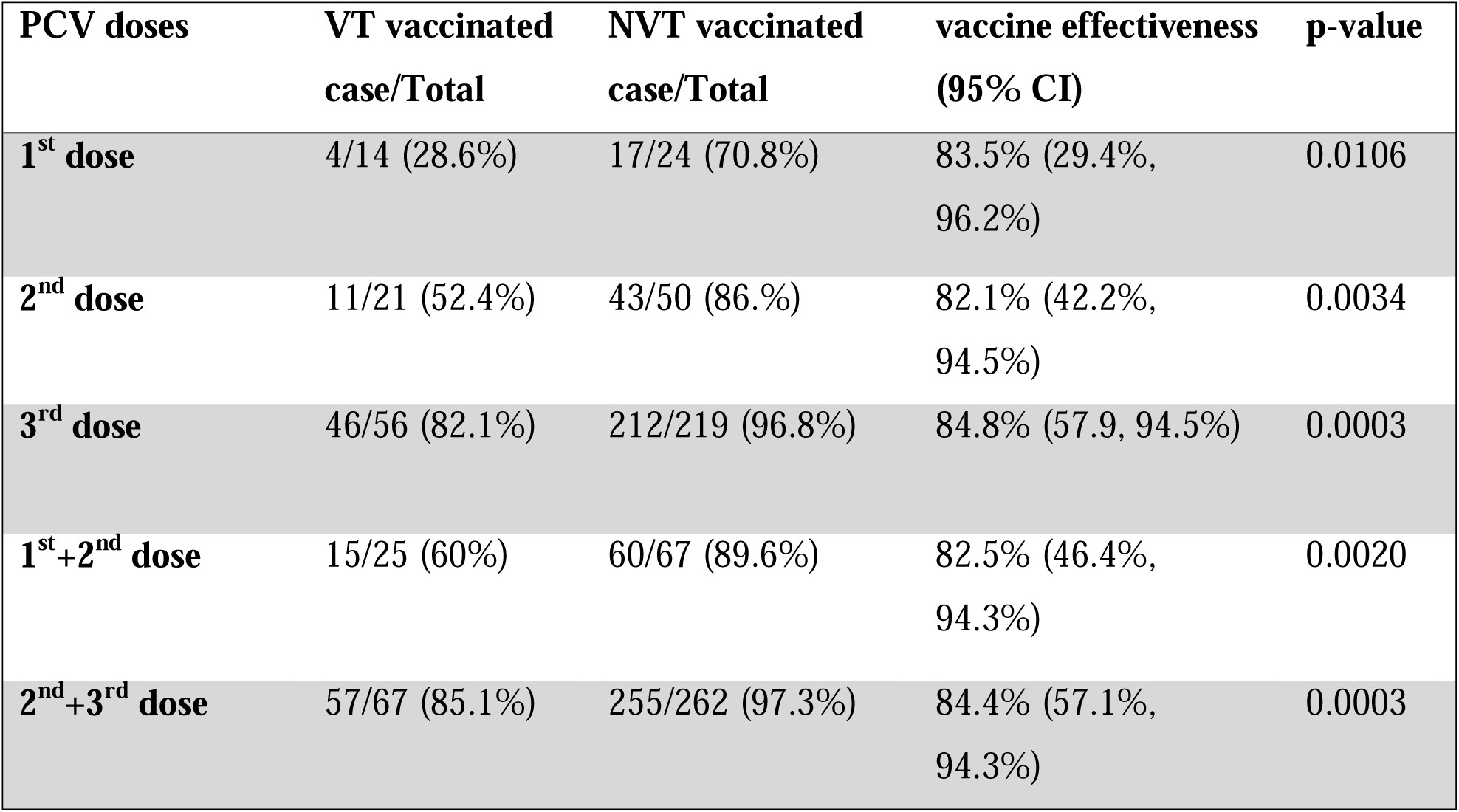
Indirect cohort analysis for PCV10 effectiveness calculation.

## DISCUSSION

Otitis media continues to be one of the most prevalent pediatric infections in Bangladesh, and a common cause of antibiotic prescriptions. This study, the largest case series of OM from Bangladesh - a country with a population of 170 million, examined the impact of PCV10 on pneumococcal OM. Specifically, it assessed changes in serotype distribution, duration of ear discharge, age distribution, and antibiotic susceptibility, allowing for the calculation of PCV10’s effectiveness in preventing OM cases cause by vaccine serotypes. To our knowledge, this represents the first report from South Asia investigating the impact of PCVs on OM.

A key finding from our study is the 15.4% reduction in pneumococcal positivity among otorrhea cases over four years following the introduction of PCV10. This reduction is consistent with systematic analyses of studies evaluating the impact of PCV on OM, which have reported reductions ranging from 8% to 42.7% for PCV7, 5.6% to 84% for PCV10, and 2.2% to 68% for PCV13 [15]. Similar results were seen from studies in Vietnam, a southeast Asian country that introduced PCV10 in March 2017 and observed a ~40% reduction in pneumococcal OM cases [21].

Our data indicated a 44.1% reduction in VT OM cases, with significant decreases observed in most PCV10 serotypes, including 19F, 6B, and 14. Limited cross-protection was noted for serotype 6A, as reported in several studies [15,22]. In contrast, no significant difference was noted for serotype 1 and low case numbers precluded effective estimates for serotypes 4, 5, and 7F. As expected, PCV10 had no effect on non-vaccine-type (NVT) OM cases. An increase in certain NVT serotype’s such as 35B was observed, along with a rise in the number of serotypes post-vaccination, frequency of NVT OM cases did not change through the study period. These changes in serotype distribution did not result in differences in median age, ear discharge duration, or the antimicrobial susceptibility against five commonly used antibiotics.

An interesting trend was observed with serotype 19A, which became an increasingly common cause of invasive pneumococcal disease in some countries following the introduction of PCVs that did not include the 19A antigen [23,24]. Some studies suggested that the 19F antigen included in PCV10 could offer cross-protection for 19A [8]. Initially, we observed a reduction in the proportion of 19A cases for two years post-vaccination. Genomic sequencing of isolates could offer an understanding of the changes in the observed phenotype [25].

Using indirect cohort analysis, the vaccine efficacy of PCV10 against VT pneumococcal OM cases was calculated at 84.4%, aligning with efficacy observed in other settings, in both developed and developing countries [15,21]. We also evaluated the coverage of other pneumococcal vaccines, noting coverage rates of 41.7% for Serum Institute PCV10, 53.5% for PCV13, 54.4% for PCV15, and 64.5% for PCV20. Incorporating PCV15 or PCV20 could potentially increase the proportion of OM cases preventable by vaccination, suggesting a benefit from higher valent pneumococcal vaccines in enhancing disease coverage.

Several limitations should be considered when interpreting our results. First, tympanocentesis was not performed on children without otorrhea, which may have led to lack of detection of certain pathogens. However, since 94% of the suspected OM cases presented with otorrhea, this limitation likely had minimal, if any, impact on our analysis. Plus, the possibility of missing bacterial pathogens in this study was low as we used selective and enriched (nonselective) media specifically for pneumococcus and *H. influenzae*. Second, we did not test for viral pathogen in the specimens, which may also play a role as co-pathogens in OM cases. Third, the study was conducted at the outpatient department of a single hospital in Dhaka, the capital city of Bangladesh. It is likely that severe OM cases, especially those with draining ears, are more inclined to seek care at facilities like Bangladesh Shishu Hospital and Institute, which is equipped to handle more complex conditions. Conversely, milder cases of OM are often managed in private clinics or pharmacies. Therefore, the impact of PCV10 observed in this study might not fully represent its effect across different healthcare settings and regions within BangladeshMoreover, having only one year of pre-PCV data limited our ability to discern longer-term trends in serotype distribution pre- and post-vaccination. Finally, the lack of a population-based denominator in our surveillance data precludes incidence calculations.

Despite these limitations, our study provides robust evidence of the significant impact of PCV10 on pneumococcal OM cases due to vaccine serotypes in Bangladesh. Studies that detail OM outcomes by specific pneumococcal serotypes are exceedingly rare, making our data particularly valuable to the existing literature. This serotype-specific analysis offers a critical perspective on vaccine effects, especially given the high burden of OM. In the absence of population-based studies, the large number of cases identified through our hospital-based surveillance enabled a detailed examination of PCV10’s impact in Bangladesh. This study underscores the value of serotype-specific investigations in understanding and enhancing vaccine effectiveness in preventing serious health outcomes. Collectively, these findings underscore the importance of continued support for PCV vaccination, highlighting its positive impact on the overall well-being of children not only in Bangladesh but across the region.

## Data Availability

All data produced in the present work are contained in the manuscript.

## AUTHOR CONTRIBUTION

SKS conceived the study, provided supervision, and secured funding. HN, MHR, ShS, MI conducted the laboratory tests. SKS, MH and MJA supervised the sample collection. HN, RCD and YH performed the data analysis and generated the figures. YH, SS, HN and SKS wrote the manuscript. YH, SS, CJW and SKS reviewed the manuscript.

## SUPPLEMENTARY FIGURES

**Sup Figure 1:**
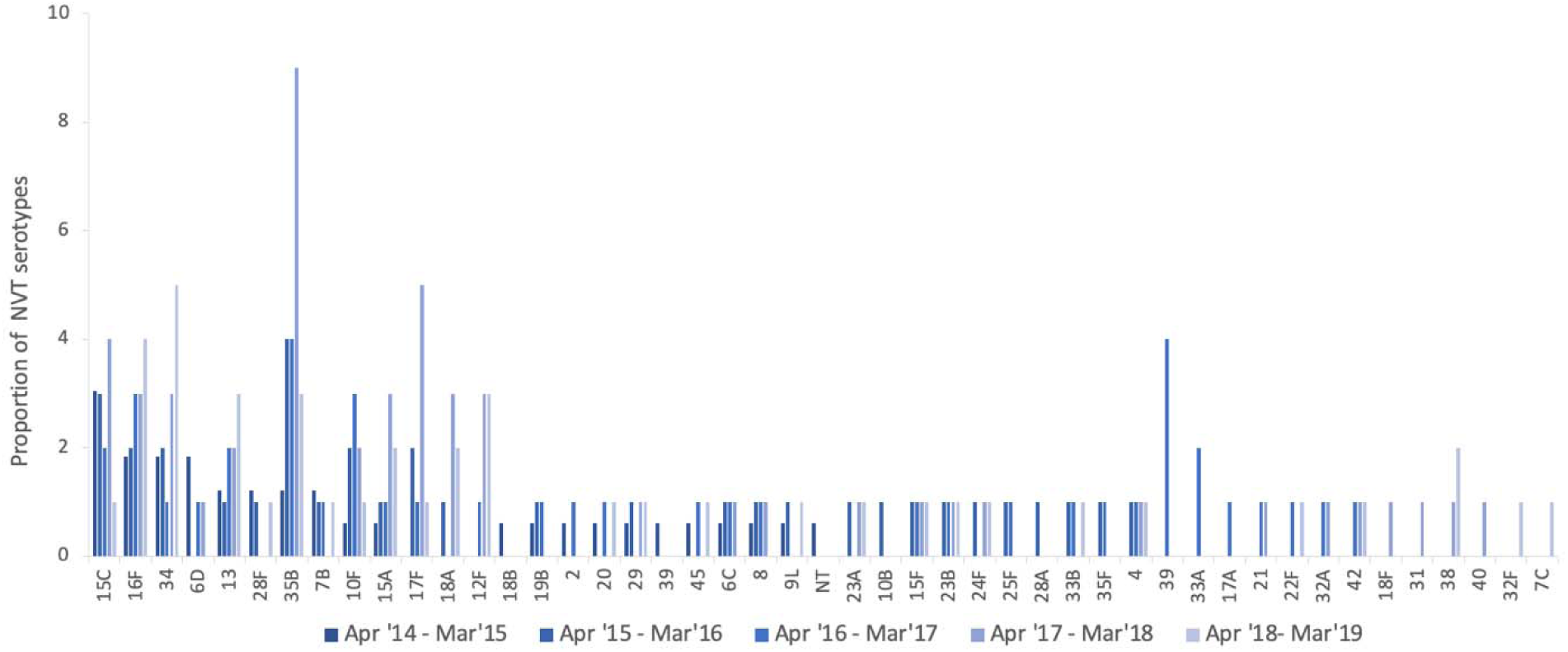
Yearly distribution of OM cases attributable to *S. pneumoniae* serotypes not covered by PCVs from April 2014 to March 2019.

**Sup Figure 2:**
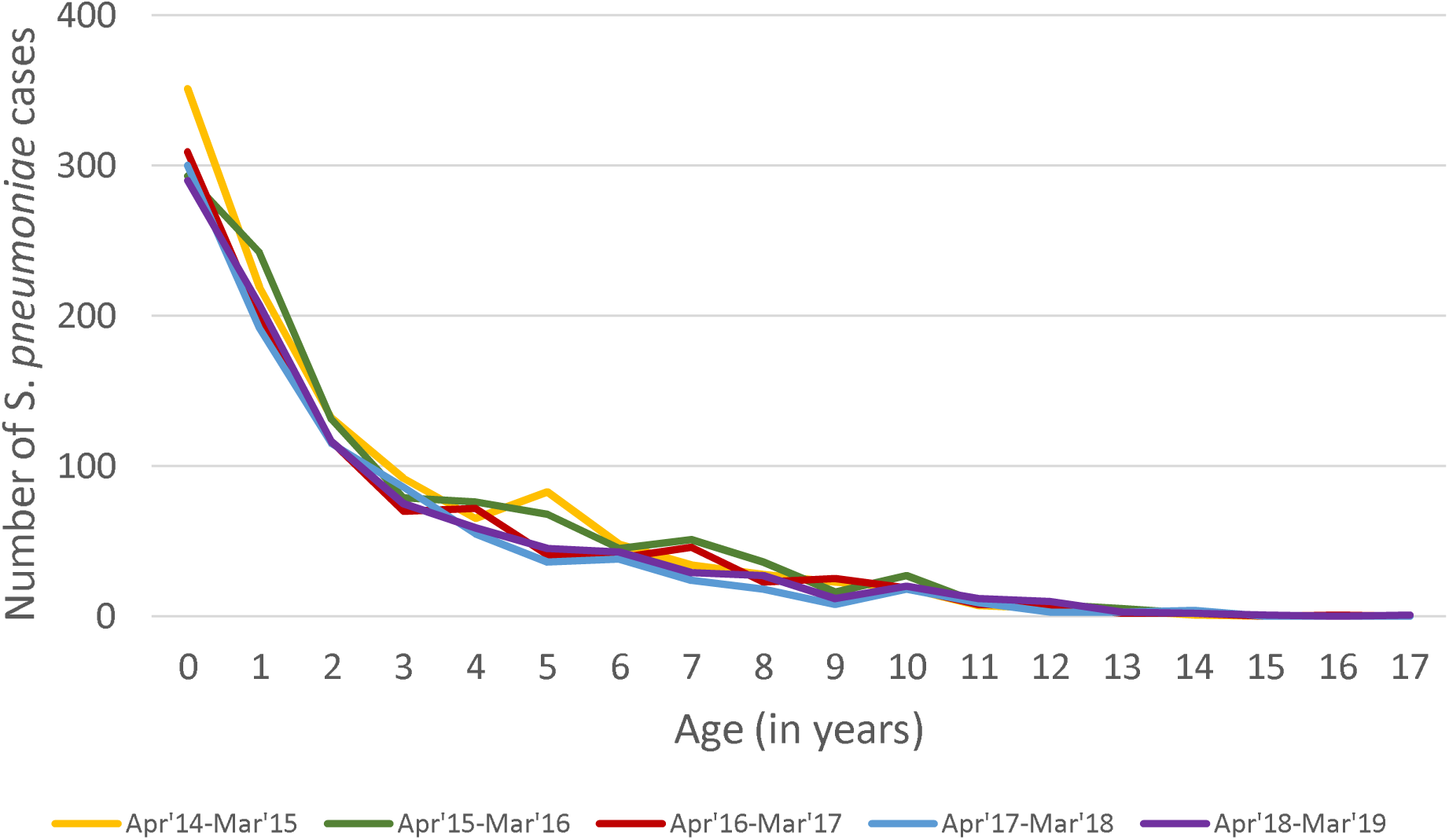
Age distribution of OM cases attributable to *S. pneumoniae* during the study period.

**Sup Figure 3:**
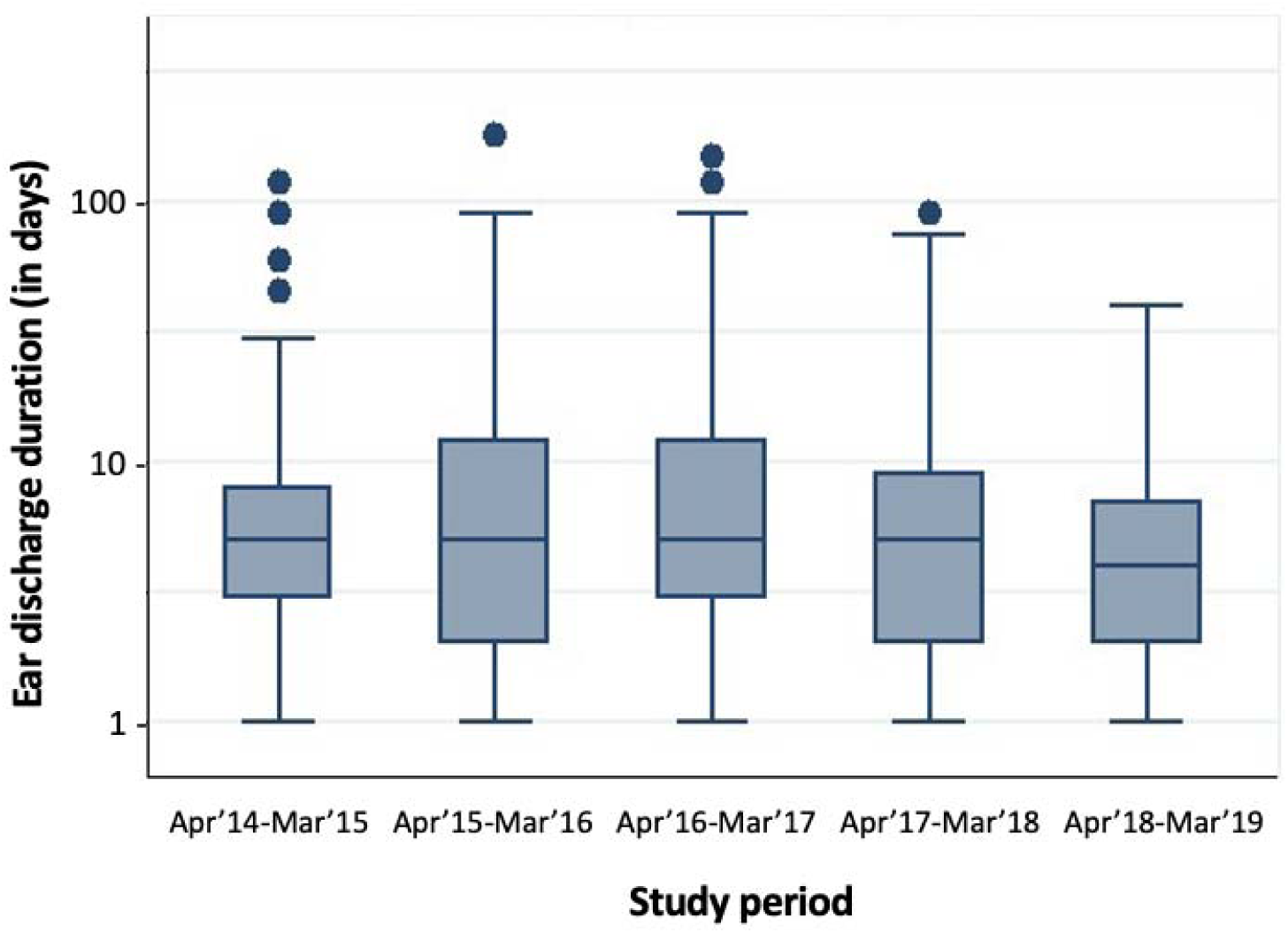
Ear discharge of OM cases attributable to *S. pneumoniae* during the study period.

**Sup Table 1:** List of all bacterial pathogens isolated from otitis media episodes detected at Bangladesh Shishu Hospital and Institute during the study period: April 2015 – March 2019 (N=3735).

| Bacterial pathogen detected | Number |
| --- | --- |
| *Haemophilus influenzae* | 418 |
| *Streptococcus pneumoniae* | 292 |
| *Staphylococcus aureus* | 308 |
| *Pseudomonas spp* | 286 |
| *Acinetobacter spp* | 43 |
| *Proteus mirabilis* | 16 |
| *Klebsiella pneumoniae* | 47 |
| *Aeromonas hydrophila* | 1 |
| *Escherichia coli* | 38 |
| *Streptococcus pyogenes* | 101 |
| *Enterobacter spp* | 27 |
| *Serratia spp* | 6 |
| *Candida* | 15 |
| *Citrobacter spp* | 4 |
| *Morganella Morganii* | 4 |
| *Streptococcus agalactiae* | 6 |
| *Klebsiella oxytoca* | 3 |
| *Proteus vulgaris* | 8 |
| *Providencia spp* | 4 |
| *Streptococcus pneumoniae* + *Haemophilus influenzae* | 171 |
| *Streptococcus pneumoniae* + *Streptococcus pyogenes* | 1 |
| *Streptococcus pneumoniae* + *Staphylococcus aureus* | 32 |
| *Streptococcus pneumoniae* + *Pseudomonas spp* | 14 |
| *Streptococcus pneumoniae* + *Acinitobacter spp* | 3 |
| *Streptococcus pneumoniae* + *Klebsiella pneumoniae* | 7 |
| *Streptococcus pneumoniae* + *Escherichia coli* | 5 |
| *Streptococcus pneumoniae* + *Enterobacter spp* | 1 |
| *Streptococcus pneumoniae* + *Providencia spp* | 3 |
| *Streptococcus pneumoniae* + *Candida* | 1 |
| *Streptococcus pneumoniae* + *Proteus mirabilis* | 4 |
| *Streptococcus pneumoniae* + *Moraxella catarrhalis* | 1 |
| *Haemophilus influenzae* + *Staphylococcus aureus* | 78 |
| *Haemophilus influenzae* + *Streptococcus agalactiae* | 1 |
| *Haemophilus influenzae* + *Moraxella catarrhalis* | 1 |
| *Haemophilus influenzae* + *Pseudomonas spp* | 12 |
| *Haemophilus influenzae* + *Streptococcus pyogenes* | 20 |
| *Haemophilus influenzae* + *Enterobacter spp* | 6 |
| *Haemophilus influenzae* + *Acinitobacter spp* | 3 |
| *Haemophilus influenzae* + *Klebsiella pneumoniae* | 7 |
| *Haemophilus influenzae* + *Escherichia coli* | 6 |
| *Haemophilus influenzae* + *Providencia spp* | 1 |
| *Haemophilus influenzae* + *Proteus mirabilis* | 1 |
| *Haemophilus influenzae* + *Streptococcus spp* | 1 |
| *Haemophilus influenzae* + *Candida* | 1 |
| *Streptococcus pyogenes* + *Acinitobacter spp* | 1 |
| *Streptococcus pyogenes* + *Klebsiella pneumoniae* | 2 |
| *Streptococcus pyogenes* + *Enterobacter spp* | 2 |
| *Streptococcus pyogenes* + *Staphylococcus aureus* | 15 |
| *Streptococcus pyogenes* + *Pseudomonas spp* | 7 |
| *Moraxella catarrhalis* + *Acinitobacter spp* | 1 |
| *Moraxella catarrhalis* + *Pseudomonas spp* | 1 |
| *Staphylococcus aureus* + *Pseudomonas spp* | 12 |
| *Staphylococcus aureus* + *Escherichia coli* | 9 |
| *Staphylococcus aureus* + *Acinetobacter* spp | 6 |
| *Staphylococcus aureus* + *Enterobacter* spp | 10 |
| *Staphylococcus aureus* + *Klebsiella pneumoniae* | 8 |
| *Staphylococcus aureus* + *Providencia* spp | 1 |
| *Staphylococcus aureus* + *Serratia* spp | 1 |
| *Staphylococcus aureus* + *Klebsiella oxytoca* | 1 |
| *Klebsiella pneumoniae* + *Escherichia coli* | 5 |
| *Klebsiella pneumoniae* + *Enterobacter* spp | 2 |
| *Klebsiella pneumoniae* + *Pseudomonas* spp | 10 |
| *Klebsiella oxytoca* + *Pseudomonas* spp | 1 |
| *E. coli* + *Pseudomonas* spp | 2 |
| *Enterobacter* spp + *Pseudomonas* spp | 2 |
| *Proteus mirabilis* + *Pseudomonas* spp | 4 |
| *Proteus mirabilis* + *Serratia* spp | 1 |
| *Acinitibacter* sp+ *Pseudomonas* spp | 1 |
| *Acinitibacter* sp+ *Citobacter* spp | 1 |
| *Streptococcus pneumoniae* + *Haemophilus influenzae* + *Acinitobacter* spp | 1 |
| *Streptococcus pneumoniae* + *Haemophilus influenzae* + *Klebsiella pneumoniae* | 1 |
| *Streptococcus pneumoniae* + *Haemophilus influenzae* + *Escherichia coli* | 2 |
| *Streptococcus pneumoniae* + *Haemophilus influenzae* + *Enterobacter* spp | 2 |
| *Streptococcus pneumoniae* + *Haemophilus influenzae* + *Providencia* spp | 1 |
| *Streptococcus pneumoniae* + *Haemophilus influenzae* + *Staphylococcus aureus* | 23 |
| *Streptococcus pneumoniae* + *Haemophilus influenzae* + *Pseudomonas* spp | 3 |
| *Streptococcus pneumoniae* + *Haemophilus influenzae* + *Streptococcus pyogenes* | 3 |
| *Streptococcus pneumoniae* + *Streptococcus pyogenes* + *Pseudomonas* spp | 2 |
| *Streptococcus pneumoniae* + *Proteus vulgaris* + *Pseudomonas* spp | 1 |
| *Streptococcus pneumoniae* + *Klebsiella pneumoniae* + *Pseudomonas* spp | 1 |
| *Streptococcus pneumoniae* + *Staphylococcus aureus* + *Acinitobacter* spp | 2 |
| *Streptococcus pneumoniae* + *Staphylococcus aureus* + *Klebsiella pneumoniae* | 1 |
| *Streptococcus pneumoniae* + *Staphylococcus aureus* + *Enterobacter* spp | 1 |
| *Streptococcus pneumoniae* + *Staphylococcus aureus* + *Proteus mirabilis* | 1 |
| *Streptococcus pneumoniae* + *Staphylococcus aureus* + *Pseudomonas* spp | 1 |
| *Haemophilus influenzae* + *Staphylococcus aureus* + *Escherichia coli* | 1 |
| *Haemophilus influenzae* + *Staphylococcus aureus* + *Acinitobacter* spp | 2 |
| *Haemophilus influenzae* + *Pseudomonas* spp + *Acinitobacter* spp | 1 |
| *Haemophilus influenzae* + *Pseudomonas* spp + *Escherichia coli* | 1 |
| *Haemophilus influenzae* + *Klebsiella pneumoniae* + *Acinitobacter* spp | 2 |
| *Haemophilus influenzae* + *Klebsiella pneumoniae* + *Enterobacter* spp | 1 |
| *Haemophilus influenzae* + *Klebsiella pneumoniae* + *Escherichia coli* | 1 |
| *Haemophilus influenzae* + *Streptococcus pyogenes* + *Acinitobacter spp* | 1 |
| *Haemophilus influenzae* + *Streptococcus pyogenes* + *Klebsiella pneumoniae* | 1 |
| *Haemophilus influenzae* + *Streptococcus pyogenes* + *Staphylococcus aureus* | 5 |
| *Haemophilus influenzae* + *Streptococcus pyogenes* + *Streptococcus agalactiae* | 1 |
| *Haemophilus influenzae* + *Staphylococcus aureus* + *Pseudomonas spp* | 2 |
| *Streptococcus pyogenes* + *Staphylococcus aureus* + *Pseudomonas spp* | 1 |
| *Streptococcus pyogenes* + *Staphylococcus aureus* + *Acinitobacter spp* | 1 |
| No bacterial pathogen detected | 1554 |
| **Total** | 3735 |

